# Circulating tumor DNA is readily detectable among Ghanaian breast cancer patients supporting non-invasive cancer genomic studies in Africa

**DOI:** 10.1101/2020.10.25.20217133

**Authors:** Samuel Terkper Ahuno, Anna-Lisa Doebley, Thomas U. Ahearn, Joel Yarney, Nicholas Titiloye, Nancy Hamel, Ernest Adjei, Joe-Nat Clegg- Lamptey, Lawrence Edusei, Baffour Awuah, Xiaoyu Song, Verne Vanderpuye, Mustapha Abubakar, Maire Duggan, Daniel Stover, Kofi Nyarko, John M S Bartlet, Francis Aitpillah, Daniel Ansong, Kevin L Gardner, Felix Andy Boateng, Anne M. Bowcock, Carlos Caldas, William D. Foulkes, Seth Wiafe, Beatrice Wiafe-Addai, Montserrat Garcia-Closas, Alexander Kwarteng, Gavin Ha, Jonine D. Figueroa, Paz Polak, on behalf of the Ghana Breast Health Study Team

## Abstract

Circulating tumor DNA (ctDNA) sequencing studies could provide novel insights into the molecular pathology of cancer in sub-Saharan Africa. ctDNA was readily detected in 15 blood samples collected in Ghana at the time of suspected breast cancer. Genomic alterations previously associated with unfavorable prognostic outcomes were observed in the majority of patients. This supports the use of liquid biopsies for diagnosis, surveillance and clinical management of breast cancer in Ghana.

---

It is well documented that there is a need to increase diversity in genomic research, particularly in African populations, where more aggressive early onset breast cancers are diagnosed, and where non-invasive methods could provide novel insights for cancer prevention and treatment. This would have both regional and global impact. Breast cancer incidence is rising in Africa and fast becoming the most common cancer on the continent where it accounts for 60% of global breast cancer deaths^1^. Technological advances in genomic and bioinformatic analysis of blood-based biomarkers offer opportunities for translational molecular oncology studies in Africa that could inform clinical-decision-making, including precision medicine treatments and new paradigms for prevention, surveillance, diagnosis and clinical management of cancer.

Whole genome sequencing of cell-free DNA (WGS-cfDNA) can identify mutations from tumor cells that shed their DNA into the bloodstream in the form of circulating tumor DNA (ctDNA) ^2-4^. ctDNA studies have been investigated in mostly European ancestry populations for possible relevance of cancer diagnosis, screening, early detection, tumor classification, and monitoring responsiveness to treatments. However, with increased investments in emerging technologies and bioinformatics, similar such studies could facilitate a quantum leap forward for molecular oncology and precision medicine in Africa^5-7^. To establish proof of concept, we used samples from Ghana, where organized mammography screening programs are lacking and late presentation is common, with the majority (62%) of breast cancer patients diagnosed with tumors >5cm^8^. Knowledge about the genomic landscape of tumors among women in Africa is limited; therefore, we tested whether ctDNA is readily detectable in Ghanaian breast cancer patients. We hypothesized that WGS-cfDNA in African women with breast cancer could reveal somatic alterations^9^ of potential clinical relevance and provide new genomic insights.

We selected 15 breast cancer patients with duplicate plasma samples collected from the Ghana Breast Health Study (GBHS)^9^. The GBHS is a population-based case-control study which recruited over 1200 incident breast cancer cases and collected samples at the time cancer was suspected. Cases were recruited from 2013-2015 from three hospital sites in the cities of Accra and Kumasi, Ghana. Demographics and tumor characteristics were similar to the overall patient population, with a median age at diagnosis of 49.5 (IQR 44.3-57.8) years. Nine patients (60%) presenting with tumors larger than 5cm, and the remaining presenting with tumors 2 to 5cm^10^. Nine (75%) of 12 patients with pathologic grade data were poorly differentiated. For three (3, 20%) patients, grade information was not available, and no tumor was classified as well-differentiated. Stage of breast cancer was not available; however, it is well documented that these patients are more frequently diagnosed with tumors with more advanced disease^10^.

Immunohistochemical (IHC) staining was available for the majority of patients and was used to classify proxies of molecular subtypes (Figure 1): Luminal A (ER+/PR+/HER2-, n=6, 40%), Luminal B (ER+ and or/PR+/HER2+, n=2, 13%), HER2-enriched (ER−/PR−/HER2+, n=2, 13%) and triple negative breast cancer (TNBC)/basal (ER-/PR-/HER2-, n=4, 27%)^11^ and indeterminate (ER missing/PR-/HER2-, n=1, 7%). cfDNA extraction and WGS at both 30x (high depth) and 0.1x (low depth) was conducted by the Broad Institute Genomic Services. The ichorCNA^12^ software was used on Next Generation Sequencing (NGS) read counts to determine the ctDNA fraction of total cfDNA and copy number alteration profiles (see Supplementary Methods for more details).

**Figure 1.**
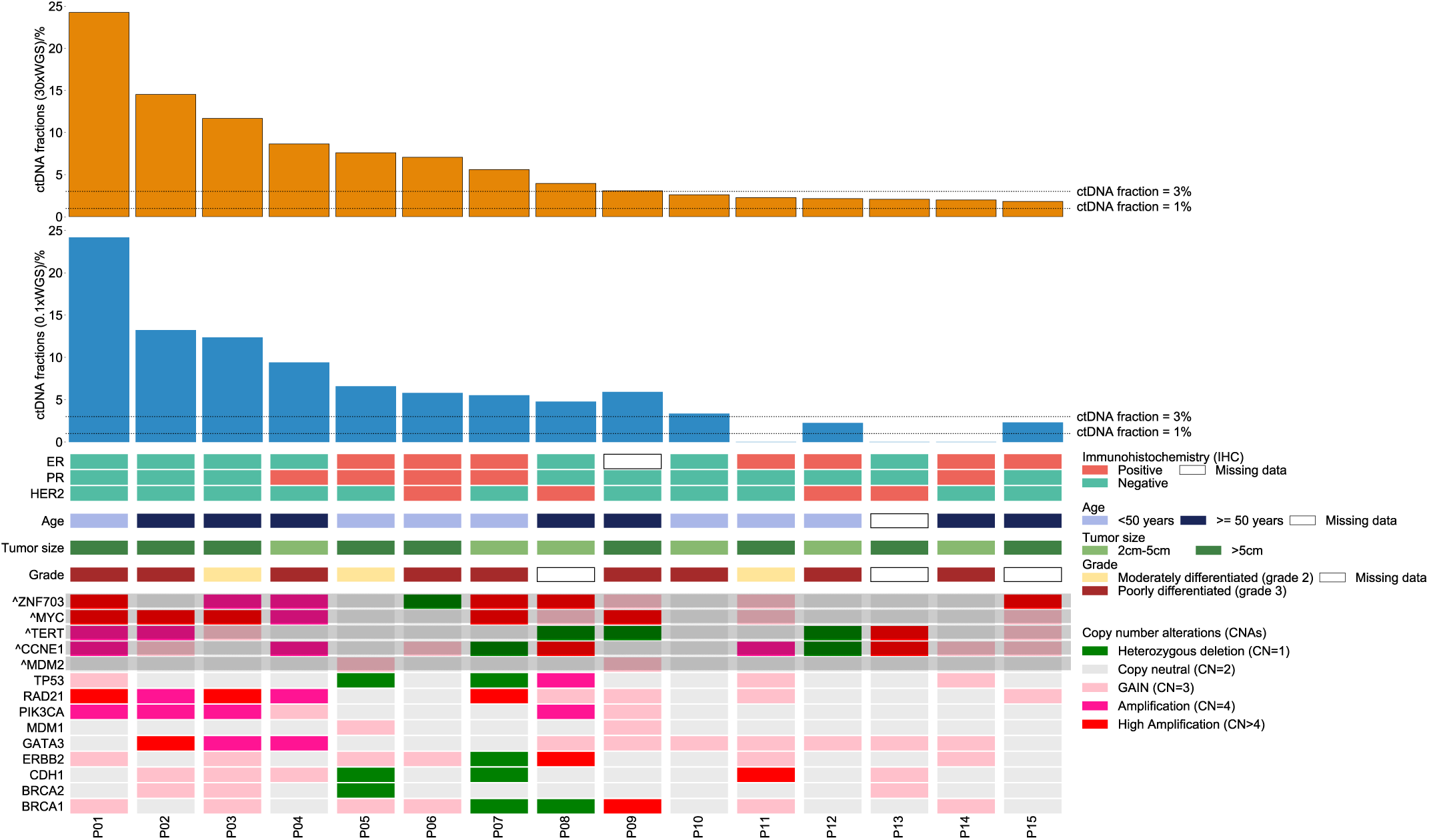
Co-mutation plot showing association between clinical information and genomic characteristics. Patients are represented by the columns ordered by decreasing ctDNA fraction. The top two rows show bar plots of tumor fractions estimated with ichorCNA from cfDNA sequenced with 30x WGS-cfDNA (gold bars) and 0.1x WGS-cfDNA (blue bars) (n=15 patients). The horizontal line across the bar plot shows the limit of detection of ichorCNA for 0.1x (ctDNA fraction= 3%) and a threshold for 30x (ctDNA fraction=1%) for the detection of ctDNA. Immunohistochemical stains for ER, PR and HER2+, age and tumor size classification are presented. Copy number gain and loss of selected genes exhibiting a high frequency of deletion or amplification in breast cancers from TCGA^13^ are shown in the bottom panel. Genes with the caret symbol (^) and shaded show copy number alterations that are significantly higher among breast cancers of African-Ancestry than in European-Ancestry patients ^13^.

The 30x WGS-cfDNA analysis indicated that all 15 breast cancer patients had at least 1% of the total cfDNA corresponded to ctDNA (median [IQR] 3.96% [2.22%-8.13%]). The tumor fractions in TNBC samples were significantly higher than the other subtypes (p=0.04; two-sided Wilcoxon rank sum test). A comparison of the estimated ctDNA fraction from the 30x cfDNA-WGS with the 0.1x WGS-cfDNA showed high concordance between estimated ctDNA fractions (Pearson r = 0.9). Using 0.1x WGS-cfDNA, a minimum of 1% ctDNA fraction was detected in 12 (80%) (95% CI 52%-96%, Figure 1).

Copy number profiling showed extensive amplification and deletion of multiple chromosomal regions (Figure 2 and Supplementary Figure 1) including those with oncogenes and tumor suppressor genes associated with breast cancer (Figure 1)^13^. We observed a high frequency (>50%) of copy number gain in three out of the five regions and potential target genes for the amplification (chr8p11-12 [ZNF703] n=8, 53.3%; chr8q24.2 [MYC] n=9, 60%; chr19q12 [CCNE1] n=9, 60%), which were previously reported to be amplified at higher frequency in patients of African-American (AA) ancestry compared to those of European-American (EA) origin in TCGA datasets^13^. Chr8p11-12 amplification is increased more than two-fold in breast tumors of AA compared to EA patients^13^ and we observed high-level amplification (more than four copies) of chr8p11-12 in four (27%, 95% CI 8%-55%) Ghanaian samples. Eight patients (53%, 95% CI 27%-79%) showed gain of at least one copy of this region that contains oncogenes ZNF703 and FGFR1 (Figure 2).

**Figure 2:**
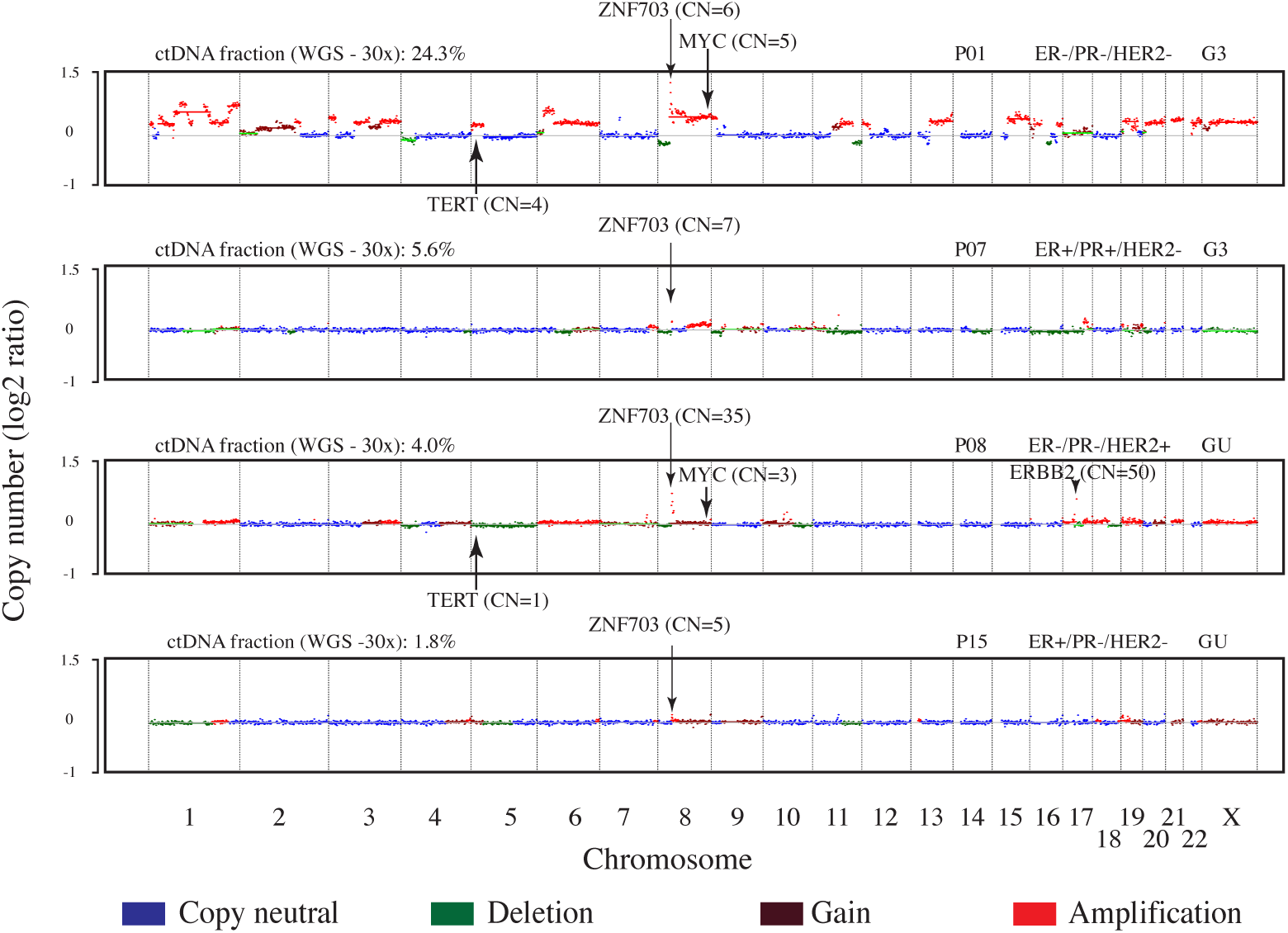
Genome-wide copy number profiles of 4 patients from cfDNA sequenced at 30x WGS. The x-axis represents chromosomes (chr1-22:X) and y-axis shows log2 copy number. Blue represents copy neutral, i.e. chromosomal loci with standard 2 copies (such as chr3, chr4, & chr5 in P07). Deletions of genomic loci (ie. chr4p in P01 & P08) are shown in green. Gains (3-4 copies) and amplifications (>4 copies) of chromosomal loci (such as chr8p12 containing ZNF703 genes in P01, P07, P08 & P15) are shown in brown and red colors, respectively. P08 has amplification of the chr17 locus containing ERBB2. Arrows indicate the gene and estimated copy number (CN) of the predicted segment by ichorCNA. GU-Grade Unknown, G2-moderately differentiated, G3-poorly differentiated, WGS-whole genome sequencing, CN-copy number, ER-estrogen receptor, PR-progesterone receptor, HER2 – human epidermal growth factor receptor 2, ctDNA-circulating-tumor DNA.

Local amplification of chr8p11-12 has been associated with luminal B tumors^14,15^ late recurrence^16^ in women of European ancestry. The amplification is associated with worse outcome than patients with no change in copy number even after adjustment of stage, grade and cancer subtype^17^. In our study, the tumors of patients with high amplification of chr8p11-12 regions were observed across the different IHC defined molecular subtypes of breast cancer. Data support ZNF703 as a driver gene that is hypothesized to be relevant in the etiology of breast cancer through regulation of both mammary basal and luminal progenitors^15^. ZNF703 amplification leads to increase in ZNF703 expression levels, which have been associated with decrease in overall survival for ER positive and luminal B patients^18^. Other potential drivers from chr8p11-12 include FGFR1 amplification within the chr8p11-12 region, which has been associated with worse outcomes in luminal A breast cancers^14^ as well as resistance to endocrine therapy^19^.

Another common high-level amplification event in our Ghana patient samples was at chr8q24, a region that contains MYC. Nine patients (60%) had at least gain (CN>= 3) of chromosomal locus chr8q24 which contains MYC. This amplification is also enriched in AA^13^ and has been linked to poor prognosis^20^. An increase in copy number was reported in 70-80% of TNBC patients^21^ and our data was consistent with this, as we observed three (75%) of four TNBC patients in our series harboring high-level amplification of this region.

Interestingly, all patients with levels of ctDNA higher than >10% were TNBC cases (3 of 3), and showed similar copy number patterns to TNBC diagnosed among patients of European ancestry^22^. These included loci harboring the two telomerase genes TERC and TERT, implicating upregulation of telomerase activity in breast oncogenesis^23^.

Lastly, of the four patients classified as HER2+ based on IHC, two had increased in copy number regions that included ERBB2 (50 and 3 copies for patients P06 and P08 respectively), consistent with data showing classification of HER2 status was variable for different quantitative assays ^24^.

Our study shows the depth of genomic information that can be obtained by interrogating WGS-cfDNA from plasma samples of breast cancer patients in Ghana. This can reveal copy number alterations and provide more detailed genomic insights into breast cancer in this and other populations with limited resources. Our observation of key alterations having similar frequencies in Ghanaian and AA breast cancer patients suggests that ctDNA may be a useful method for comparative genomic studies of breast cancer as well as for other cancers. In particular targeted assays for the detection of high copy number of ZNF703/FGFR1 and MYC would increase the sensitivity of their detection in the low fractions of ctDNA circulating in the blood. As this amplification is established to be associated with late stage tumors^17^, future research is needed to determine factors associated with its higher frequency, and in particular whether it is due to presentation at a late cancer stage or is present due to other genetic, environmental or lifestyle associated factors.

On the African continent, cancer incidence is rising, mortality rates are high and access to molecular pathology is limited. Non-invasive methods for molecular tumor subtyping could be transformative. The rapid advancement of sequencing technologies and the estimated ctDNA fraction levels (>1%) in this sample of Ghanaian patients in this study indicate that the exploration of alternative methods such as methylation-based cfMedip-seq and panel testing are warranted. These could have high sensitivity (for at least 0.1% ctDNA fraction) and be suitable for patients at the time of diagnosis^2,4,12^. Moreover, the ability to detect tumor with 0.1x WGS-cfDNA support additional advances for sample collection methods (such as a finger prick blood test^25^). New long read sequencing technologies (such as Nanopore sequencing) would allow more facile translation of cancer genomic data to be used at the point of care. Hence, our study shows that an analysis of ctDNA could be a tool for future molecular oncology studies in Africa that merits further investment for cancer etiology, surveillance, molecular oncology and clinical trial studies that are urgently needed to improve cancer survival in Africa.

## Data Availability

Raw sequencing data will be deposit in dbGaP and can be used for breast cancer research and other adult disease only.

## Acknowledgement

The authors would like to thank the patients for donating samples to the GBHS study. We would also like to thank Jonathan B Reichel (University of Washington) for helpful comments in preparation of the manuscript. This study was supported in part by a Komen Leadership award to WDF (SAC110008). GH is supported by NCI K22 CA237746. PP is supported by the Jimmy V foundation scholar award. JDF acknowledges personal funding on molecular subtypes of breast cancer from the Wellcome Trust (207800/Z/17/Z) and MRC (MR/S015027/1).

The authors acknowledge the research contributions of the Cancer Genomics Research Laboratory for their expertise, execution, and support of this research in the areas of project planning, wet laboratory processing of specimens. This project has been funded in whole or in part with Federal funds from the National Cancer Institute, National Institutes of Health, under NCI Contract No. 75N910D00024. The content of this publication does not necessarily reflect the views or policies of the Department of Health and Human Services, nor does mention of trade names, commercial products, or organizations imply endorsement by the U.S. Government.

## Author contribution

Conception and design of the study: P.P, W.D.F, J.D.F. and G.H. The main bioinformatic and computational analysis: S.T.A. Assisting in the computational work: A.L.D, G.H., and P.P. Data and sample collection: L.E, N.T, E.A, J.N.C.L, J.Y, B.W.A, B.A, V.V, M.D, S.W, K.N, F.A, D.A and Ghana Breast Health Study team. Sample selection and phenotype data: T.U.H. and M.G.C. Pathology review: L.E., N.T., E.A., M.D. Statistical analysis: S.T.A, P.P, G.H., X.S, and J.D.F. Interpretation of data: all authors. Drafting of the paper: S.T.A, P.P, W.D.F, J.D.F., N.H. and G.H. Revised work and provided important intellectual content: all authors. Final approval of the paper: all authors. P.P, G.H. and J.D.F. equally supervised this work.

## Ghana Breast Health Study team

Ghana Statistical Service, Accra, Ghana: Dr Robertson Adjei and Dr Lucy Afriyie. Korle Bu Teaching Hospital, Accra, Ghana: Professor Dr Anthony Adjei, Dr Florence Dedey, Victoria Okyne, Naomi Ohene Oti, Evelyn Tay (deceased), Dr Adu-Aryee, Angela Kenu and Obed Ekpedzor. Komfo Anoyke Teaching Hospital, Kumasi, Ghana: Marion Alcpaloo, Isaac Boakye, Bernard Arhin, Emmanuel Assimah, Samuel Ka-chungu, Dr Joseph Oppong and Dr Ernest Osei-Bonsu. Peace and Love Hospital, Kumasi, Ghana: Prof Margaret Frempong, Emma Brew Abaidoo, Bridget Nortey Mensah, Samuel Amanama, Prince Agyapong, Debora Boateng, Ansong Thomas Agyei, Richard Opoku and Kofi Owusu Gyimah. New York-Presbyterian/Weill Cornell Medical Center, NY, USA: Dr Lisa Newman. National Cancer Institute, Bethesda, MD, USA: Louise A. Brinton, Maya Palakal and Jake Thistle. Westat, Inc.: Michelle Brotzman, Shelley Niwa, Usha Singh and Ann Truelove. University of Ghana: Prof Richard Biritwum.

## Ethical approval

The study was approved by the Special Studies Institutional Review Board of the National Cancer Institute (Rockville, MD), the Ghana Health Service Ethical Review Committee and institutional review boards at the Noguchi Memorial Institute for Medical Research (Accra, Ghana), the Kwame Nkrumah University of Science and Technology (Kumasi, Ghana), the School of Medical Sciences at Komfo Anokye Teaching Hospital (Kumasi, Ghana) and Westat (Rockville, MD). All participants provided written informed consent.

## Code availability

Code for ichorCNA v0.2.0 used to analyze 0.1x WGS-cfDNA data can be accessed at https://github.com/broadinstitute/ichorCNA. Analysis of 30x WGS-cfDNA data used an updated version of ichorCNA and can be accessed at https://github.com/GavinHaLab/ichorCNA [commit b2bbce0, June 22, 2020].

