## Supplementary Methods for "Circulating tumor DNA is readily detectable among Ghanaian breast cancer patients supporting non-invasive cancer genomic studies in Africa"

This pdf file includes;

- Materials and Methods
- Supplementary Figure
- References
- Appendix

**Supplementary Figure**
Figure 1. Genome-wide copy number profiles of all 15 patients from cfDNA sequenced at 30x Whole Genome Sequencing (WGS) ………………………………………………………….……. 7

### Materials and Methods

##### 1.1. Clinical specimens - tumor biopsy

This project is based on a case-control study of breast cancer conducted in collaboration with three hospitals in Ghana responsible for treating most breast cancers in the country which has been previously described;[1,2] Korle Bu Teaching Hospital (KBTH) in Accra and Komfo Anokye Teaching Hospital (KATH) and Peace and Love Hospital (PLH) in Kumasi. Eligible cases in this study were women 18-74 years of age who were residents of defined catchment areas surrounding Kumasi and Accra for at least one year's time and who in the preceding year had a lump suspected to be cancer that resulted in either referral for a biopsy at the study hospitals (KBTH, KATH, and PLH) or for clinical care. In addition to extracting information from medical records into standardized case abstract forms, study pathologists (Dr. Duggan) performed a centralized histopathology review of H&E sections from biopsy formalin-fixed paraffin embedded (FFPE) blocks sent to the US (~75% of malignant cases).

##### 1.2. Clinical specimens - blood samples

Venous blood samples were collected using a 10ml purple top EDTA tube at the time of recruitment, which was usually the time of suspected breast cancer. Purple top tubes were centrifuged and processed within two hours of collection to avoid red blood cell (RBC) contamination of the separated plasma layer. Each collected purple-top tube was kept at room temperature (18-25ºC or 64-77ºF) and processed for a minimum of 15 minutes at 1500g / Relative Centrifugal Force (RCF). The top plasma layer was separated from buffy coat, and red blood cells. Three 1.8 ml plasma aliquots were stored using cryovials. Processed plasma specimens were stored at -80°C freezers. cfDNA was extracted from 2 aliquots of 1.8 ml plasma samples and sequenced at low depth (0.1x ULPS) and at higher depth (30x Whole Genome Sequencing, WGS).

##### 1.3. Tumor characterization (tumor grade, size, and Immunohistochemistry)

FFPE blocks received from Ghana were re-embedded prior to obtaining 5µm H&E sections for pathology review by study pathologists using a standardized form to capture the quality of the specimens and histopathology diagnosis and grade of tumors (See Appendix A). IHC data was based on assessment in Ghana as recorded on case abstract forms. A concordance study comparing IHC staining results by Ghanaian pathologists from diagnostic blocks in Ghana and by an NCI pathologist using the study blocks at NCI (i.e., different FFPE blocks from the same patient) showed good agreement for ER (79% agreement based on 87 cases) and HER2 (78% based on 76 cases); however, the agreement for PR was lower (65% based on 86 cases) as previously described[2].

Tumor size was based on clinical assessment as previously described[1]. At the time of biopsy, data were recorded by nurses and physicians on the presenting symptoms, number of lumps/masses present, and the approximate size of the lumps/masses from which biopsies were obtained.

##### 1.4. Library construction and sequencing of cfDNA

cfDNA was sequenced at the Broad Institute's Genomics Services using 5-10ml blood, 4-6ml plasma, or 2-20ng cfDNA. The process utilizes the DSP Circulating DNA kit from Qiagen to extract cell-free DNA from plasma and elute into 40-80uL of the re-suspension buffer using the Qiagen Circulating DNA kit on the QIAsymphony liquid handling system [3,4]. Extracted cell-free DNA is stored frozen at -20 C until ready for further processing. CfDNA were sequenced on Illumina next-generation sequencing platform.

##### 1.5. Next Generation Sequencing (NGS) data preprocessing

Raw reads in the form of fastq files were preprocessed using the GATK best practices guidelines for NGS data (Figure 1). This involves mapping of raw reads to human reference genome (Hg19/GRCh37) using Burrows-Wheeler Aligner (BWA-MEM) algorithms [5] to generate a Sequence Alignment/Map (SAM) file of mapped reads. Sam files were converted to a binary alignment format (BAM) file of aligned reads [6].

Duplicate reads from Polymerase Chain Reaction (PCR) during the library preparation step of sequencing were removed with "gatk markduplicates" to allow us to confidently detect which DNA segments are amplified or delete due to cancer (biological effect and not technical) [7]. The base quality score recalibration (BQSR) step which employs machine learning algorithms (GATK BaseRecalibrator tool) to model the systematic errors from the sequencing machine and correct them (with gatk ApplyBQSR tool) were then carried out as final step for the GATK best practices for data preprocessing after which an analysis-ready-bam file is generated for tumor fraction estimation, ploidy, and copy number alterations profiles with ichorCNA software

All the data preprocessing steps were carried out in Terra (https://app.terra.bio/), a cloud-based analysis platform developed by the Broad Institute in collaboration with google cloud, with publicly available workflows.

##### 1.6. Tumor fraction detection and copy number profiles prediction

To estimate the tumor fraction (cell-free DNA coming from neoplastic cells) and copy number alterations, the ichorCNA software was used [4]. The software accepts an analysis-ready-bam as input and simultaneously estimates tumor fractions and copy number profiles through a three-step process 1) dividing the genome into non-overlapping windows of specific length and computing read coverage within each bin, 2) read normalization; correcting for Guanine-Cytosine content, replication timing, mappability of reads, 3) tumor fraction estimation and CNA prediction; implemented using Hidden Markov Models (HMM) segmentation algorithm to find neighboring bins as genomic regions that are amplified or deleted together and the expectation-maximization algorithms to estimate the parameters of the model.

For 0.1x ULPS analysis, ichorCNA (v0.2.0) release version (https://github.com/broadinstitute/ichorCNA) was used with default settings (https://github.com/broadinstitute/ichorCNA/blob/master/scripts/snakemake/config/config.yaml).

Briefly, the configuration settings are the following:
A 1MB bin size was used to compute coverage. The following resources, Guanine+Cytosine (GC) score, and mappability all in 1MB wig file format for hg19 were used to normalize reads when running ichorCNA. Centomere files (resource) based on UCSC (GRCh37.p13) were downloaded from UCSC and used to mask repetitive sequences in the human genome that the aligner may have erroneously mapped. A panel of normals (PoN) from 27 healthy donors provided in the ichorCNA package was used. The threshold applied for minimum mapping score was 0.75. To control segmentation, which will ultimately affect sensitivity, the transition probabilities were set as ichorCNA_txnE=0.99 and ichorCNA_txnStrength=100. The non-tumor fraction parameter restart values of 0.5,0.6,0.7,0.8,0.9,0.95 were used. Tumor ploidy restart values of 2 and 3 were used. Clonality settings were used for subclonal copy number 1 and 3. Maximum copy number of 5 was set and a Student's-t likelihood model was used. Solutions for each parameter restart were ranked by the log-likelihood value, and only solutions were included if the following criteria were met: Maximum fraction of genome accounted as subclonal was <0.5, the maximum fraction of genome in subclone was <0.7. Then, the optimal solution was selected from the remaining solution with the highest log-likelihood. Manual inspection of these solutions were performed to confirm the results.

In the 30x WGS analysis, an updated version of ichorCNA (https://github.com/GavinHaLab/ichorCNA) was used with custom configurations to analyze deeper WGS data and to increase the sensitivity needed to be able to detect low tumor fractions. Parameters used for analysis are stored in config files and are publicly available on github (https://github.com/sahuno/cfDNA_Ghana_pilot_GBHS/tree/master/scripts/ichorCNA_configs/ WGS_30x_GBHS_ichorCNA_config).

The key distinguishing steps here are that;
1) mappability was accounted for
2) ichorCNA_txnStrength was decreased from default value of 10000 to 100 to employ more segments leading to higher sensitivity.
3 ) The ichorCNA_txnE used was 0.99 to increase sensitivity.
4) The –minSegmentBins was reduced from 50 to 20
4) The –altFracThreshold was reduced 0.05 to 0.01

##### 1.7. Annotation of somatic copy number variants

To annotate copy number alterations, Ensembl genes[8] (human reference genome GRCh37.p13) with coordinates (Chromosomes 1:22, X, Y, without patches) were retrieved from Ensembl legacy website http://grch37.ensembl.org/biomart/martview/2c80f6edb70f976fa9dbcdc5127d8cf0 (accessed date; Friday, February 14, 2020) programmatically with R package Biomart[8].

The following attributes were used to query reference genes from Ensembl, Gene stable ID, Chromosome/scaffold name, Gene start (bp), Strand (+/-), Gene end (bp), Gene name, Human Genome Organization (HUGO) Gene Nomenclature Committee (HGNC) symbol, Gene type. The queried Ensembl gene list with coordinates and metadata and copy number segments were converted to Genomic ranges object using the `GenomicFeatures` package to allow complete overlap between unique genes and copy number segments. The Genomic ranges object is a type of object in R used to conveniently store gene information (chromosome, start position, end position, strand) and other metadata of the gene (gene name, gene ID, HGNC symbol, etc)

The frequency of copy number changes was calculated for a subset of genes known to be recurrently altered in chromosomal segments extracted from TCGA (cbioportal, pan cancer breast study) sorted according to GISTIC scores/q-value. GISTIC (Genomic Identification of Significant Targets in Cancer) is an algorithm for detecting possible cancer driver genes in copy number profiles by evaluating the frequency and amplitude of the events (loss and/or gain of genomic regions) [9]. Hence the associated GISTIC q-value and score was the metric used for accepting possible drivers [9].

Also, genes differentially altered among African Americans and White were compared in our cohort. For simplicity, the HGNC symbols of annotated genes were used throughout the report of the analysis also given that they are unique and meaningful naming schemes for known human genes [10].

##### 1.8. Statistical analysis

Descriptive statistics were calculated for variables of clinical information. Median interquartile (IQR) of ctDNA fractions and ploidy were computed and compared with clinicopathological information such as age, grade, immunohistochemistry stains (IHC), hormonal receptor expression, and subtypes.

### Supplementary Figure

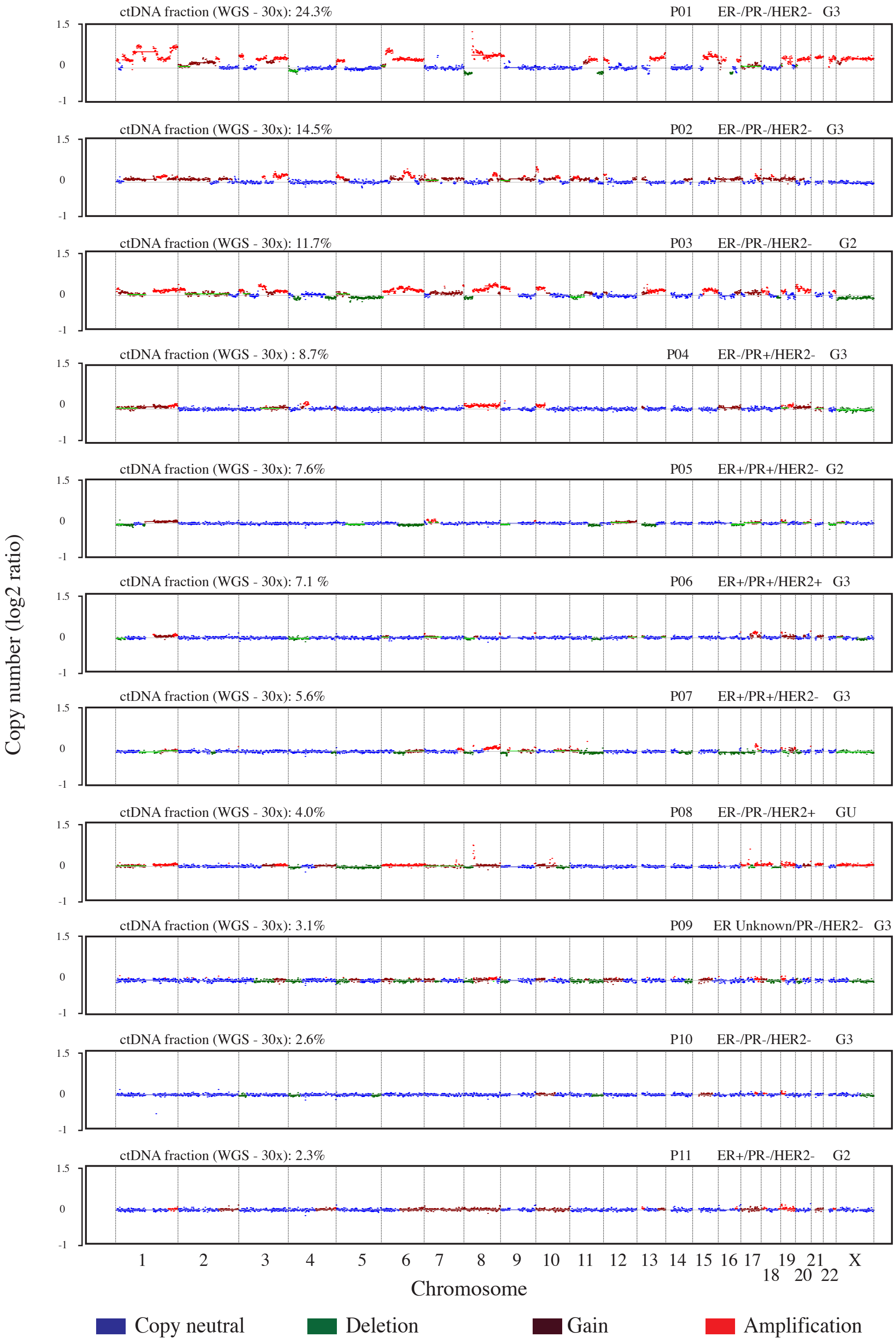

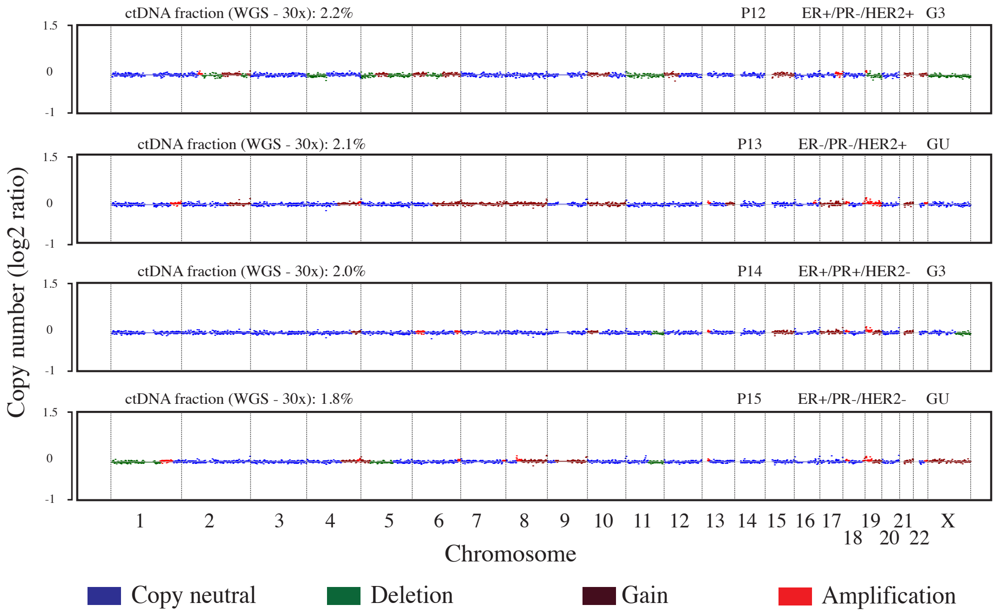

Supplementary Figure 1: Genome-wide copy number profiles of all 15 patients from cfDNA sequenced at 30x Whole Genome Sequencing (WGS). Genome-wide copy number profiles of the fifteen (15) cases arranged in descending order of tumor fractions. The y-axis corresponds to log2 copy ratio estimated by ichorCNA and x-axis are chromosomes (chr1-22 and chrX). Patient IDS, tumor grade, Immunohistochemistry stains, tumor fractions are on top of each plot. GU- Grade Unknown, G2- moderately differentiated, G3-poorly differentiated, WGS- whole genome sequencing, CN-copy number, ER- estrogen receptor, PR-progesterone receptor, HER2 – human epidermal growth factor receptor 2, ctDNA- circulating-tumor DNA

### APPENDIX: H&E Pathology review form

BSI ID: TBR______________________________ REVIEWER:_____________________ DATE REVIEWED:______________________

| Technical Preparation | Tissue Adequacy | Tumor Adequacy |
|---|---|---|
| 1a. Satisfactory ........ 1 [Go to 2a]<br>Limited ........ 2 [Go to 1b]<br>Unsatisfactory ........ 3 [Go to 1b] | 2a. Satisfactory ........ 1 [Go to 3a]<br>Limited ........ 2 [Go to 2b]<br>Unsatisfactory ........ 3 [Go to 2b] | 3a. Satisfactory ........ 1<br>Limited ........ 2 [Go to 3b]<br>Unsatisfactory ........ 3 [Go to 3b] |
| 1b. Circle all that apply:<br>Missing slide ... 1 Cover slipping issues ... 4<br>Broken slide .... 2 Other ........ 5<br>Folds ........ 3 Specify ____________ | 2b. Circle all that apply:<br>Crushed (Partial) ........ 1 Fixation issues (Partial) ........ 5<br>Crushed (Complete) ... 2 Fixation issues (Complete) .... 6<br>Necrotic (Partial) ........ 3 Other ........ 7<br>Necrotic (Complete) ... 4 Specify ________________ | 3b. Circle all that apply:<br>No tumor ........ 1 Necrotic (Complete) ........ 6<br>Small tumor volume ... 2 Fixation issues (Partial) ....... 7<br>Crushed (Partial) ....... 3 Fixation issues (Complete) .. 8<br>Crushed (Complete) .. 4 Other ........ 9<br>Necrotic (Partial) ........ 5 Specify ________________ |

4a. Is Invasive cancer present: Yes ........ 1 [Go to 4b and 4c]
No ........ 2 [Go to 5a]

4b. **Invasive Diagnosis** (Circle all that apply)

| Type | Classification |
|---|---|
| Invasive carcinoma of no special type (NST) | 8500/3 |
| Pleomorphic carcinoma | 8522/3 |
| Carcinoma with osteoclast-like stromal giant cells | 8035/3 |
| Carcinoma with choriocarcinomatous features | |
| Carcinoma with melanotic features | |
| Invasive lobular carcinoma | 8520/3 |
| Classic lobular carcinoma / Pleomorphic lobular carcinoma | |
| Solid lobular carcinoma / Tubulolobular carcinoma | |
| Alveolar lobular carcinoma / Mixed lobular carcinoma | |
| Tubular carcinoma | 8211/3 |
| Cribriform carcinoma | 8201/3 |
| Mucinous carcinoma | 8480/3 |
| Carcinoma with medullary features | |
| Medullary carcinoma | 8510/3 |
| Atypical medullary carcinoma | 8513/3 |
| Invasive carcinoma NST with medullary features | 8500/3 |
| Carcinoma with apocrine differentiation | |
| Carcinoma with signet-ring-cell differentiation | |
| Invasive micropapillary carcinoma | 8507/3 |
| Metaplastic carcinoma of no special type | 8575/3 |
| Low-grade adenosquamous carcinoma | 8570/3 |
| Fibromatosis-like metaplastic carcinoma | 8572/3 |
| Squamous cell carcinoma | 8070/3 |
| Spindle cell carcinoma | 8032/3 |
| Metaplastic carcinoma with mesenchymal differentiation | |
| Chondroid differentiation | 8571/3 |
| Osseous differentiation | 8571/3 |
| Other types of mesenchymal differentiation | 8575/3 |
| Mixed metaplastic carcinoma | 8575/3 |
| Myoepithelial carcinoma | 8982/3 |
| *Epithelial-myoepithelial tumors* | |
| Adenomyoepithelioma with carcinoma | 8983/3 |
| Adenoid cystic carcinoma | 8200/3 |
| *Rare types* | |
| Carcinoma with neuroendocrine features | |
| Neuroendocrine tumor, well-differentiated | 8246/3 |
| Neuroendocrine carcinoma poorly differentiated (small cell carcinoma) | 8041/3 |
| Carcinoma with neuroendocrine differentiation | 8574/3 |
| Secretory carcinoma | 8502/3 |
| Invasive papillary carcinoma | 8503/3 |
| Acinic cell carcinoma | 8550/3 |
| Mucoepidermoid carcinoma | 8430/3 |
| Polymorphous carcinoma | 8525/3 |
| Oncocytic carcinoma | 8290/3 |
| Lipid-rich carcinoma | 8314/3 |
| Glycogen-rich clear cell carcinoma | 8315/3 |
| Sebaceous carcinoma | 8410/3 |

4c. Grade: Grade 1 .... 1 Grade 3 ........ 3 Not applicable ... 5
Grade 2 .... 2 Unable to grade ... 4

4d. Number of invasive cancer pieces: __________

4e. Number of tissue pieces present: __________

5a. Are non-invasive precursor lesion(s) present: Yes ....... 1 [Go to 5b]
No ........ 2 [Go to 6a]

5b. **Non-invasive Precursor Lesions Diagnosis** (Circle all that apply)

| Type | Classification |
|---|---|
| Precursor lesions | |
| Ductal carcinoma in situ | 8500/2 |
| Lobular neoplasia | |
| Lobular carcinoma in situ | |
| Classic lobular carcinoma in situ | 8520/2 |
| Pleomorphic lobular carcinoma in situ | 8519/2* |
| Atypical lobular hyperplasia | |
| Intraductal proliferative lesions | |
| Usual ductal hyperplasia | |
| Columnar cell lesions including flat epithelial atypia | |
| Atypical ductal hyperplasia | |
| Papillary lesions | |
| Intraductal papilloma | 8503/0 |
| Intraductal papilloma with atypical hyperplasia | 8503/0 |
| Intraductal papilloma with ductal carcinoma in situ | 8503/2* |
| Intraductal papilloma with lobular carcinoma in situ | 8520/2 |
| Intraductal papillary carcinoma | 8503/2 |
| Encapsulated papillary carcinoma | 8504/2 |
| Encapsulated papillary carcinoma with invasion | 8504/3 |
| Solid papillary carcinoma | |
| In situ | 8509/2 |
| Invasive | 8509/3 |

6a. Are benign lesions present: Yes ........ 1 [Go to 6b]
No ........ 2 [End]

6b. **Benign Lesion Diagnosis** (Circle all that apply)

| | |
|---|---|
| No pathological abnormality | 1 |
| Abscess +/- organization | 2 |
| Scar | 3 |
| Cyst/Cystically dilated duct +/- apocrine metaplasia | 4 |
| Ruptured Cyst/Cystically dilated duct +/- macrophage reaction | 5 |
| Fat necrosis | 6 |
| Stromal fibrosis | 7 |
| Stromal calcification | 8 |
| Pseudolactational changes, hyperplasia | 9 |
| Fibroadenoma | 10 |
| Sclerosing adenosis | 11 |
| Radial scar | 12 |
| Tubular adenoma | 13 |
| Intramammary lymph node | 14 |
| Granulomatous inflammation | 15 |
| Other ________________________________ | 16 |

6c. Diagnosis Not Possible ........ 1
